# A pan-European study of SARS-CoV-2 variants in wastewater under the EU Sewage Sentinel System

**DOI:** 10.1101/2021.06.11.21258756

**Authors:** Shelesh Agrawal, Laura Orschler, Selina Schubert, Kira Zachmann, Leo Heijnen, Simona Tavazzi, Bernd Manfred Gawlik, Miranda de Graaf, Gertjan Medema, Susanne Lackner

**Affiliations:** Technical University of Darmstadt, Department of Civil and Environmental Engineering Sciences, Institute IWAR, Chair of Wastewater Engineering; Darmstadt, Germany; KWR Water Research Institute; Nieuwegein, The Netherlands; European Commission, Joint Research Centre; Ispra (Va), Italy; Department of Viroscience, Erasmus Medical Center; Rotterdam, The Netherlands

## Abstract

Wastewater based surveillance employing qPCR has already shown its utility for monitoring SARS-CoV-2 at community level, and consequently the European Commission has recommended the implementation of an EU Sewage Sentinel System. However, using sequencing for the determination of genomic variants in wastewater is not fully established yet. Therefore, we focused on the sequencing analysis of SARS-CoV-2 RNA in wastewater samples collected across 20 European countries including 54 municipalities. Our results provide unprecedented insight into the abundance and the profile of the mutations associated with the variants of concerns: B.1.1.7, P.1, B.1.351 and B.1.617.2, which were present in various wastewater samples. This study shows that integrating genomic and wastewater-based epidemiology (WBE) can support the identification of variants circulating in a city at community level.

## Introduction

Undeniably, the sudden emergence of SARS-CoV-2, which has caused a global pandemic, is a significant and in many regards unprecedented threat to public health. SARS-CoV-2 rapidly resulted in a high number of people requiring hospitalization, casualties and major socio-economic disruptions, with consequences which we still do not fully oversee. Consequently, most countries have been forced to implement severe lockdown measures to ensure the physical distance between people and interrupt virus transmission (1). Overall, the high transmission rate and the rapidly evolving nature of the virus, leading to the emergence of new variants that may transmit more readily and evade the immune response, raise broad concerns about SARS-CoV-2 (2–4).

The current phase of the COVD-19 pandemic is shaping into an era of genomic surveillance to track the genomic changes in the SARS-CoV-2 virus, which belongs to the family *Coronaviridae*, genus *Betacoronavirus*. According to PANGO lineages as of now, 1503 SARS-CoV-2 variants are known since the initial detection of SARS-CoV-2 by sequencing (5). In the last few months, genomic epidemiology, the analysis of genome sequences, has revealed some fast-spreading and highly virulent SARS-CoV-2 variants (3, 6, 7), making them variants of concern (VOC) (8, 9). This development underlines the importance of sequencing analyses. Although the European Commission recommended to sequence 5-10% of the SARS-CoV-2 positive patient samples, as of 22 January 2021 (10), most Member States are below this recommended sequencing target (11).

Wastewater-based epidemiology (WBE) is an emerging paradigm for monitoring the circulation of SARS-CoV-2 in a community. Several research groups across the globe have shown that WBE provides additional information about the dynamics of SARS-CoV-2 at community level (12–17). WBE efforts have primarily focused on quantitative polymerase chain reaction (qPCR), determining the titers of SARS-CoV-2 in the sewage and its correlation to the reported number of SARS-CoV-2 positive cases. At present, very few studies are available that looked for the combined potential of genomic epidemiology and WBE to determine the SARS-CoV-2 genomic variants circulating in a specific region (18–21). This pan-European study, which has been conducted under the umbrella of the EU Sewage Sentinel System for SARS-CoV-2 (22), a direct result of what is called “The HERA Incubator” (23), assesses whether next-generation sequencing (NGS) of wastewater samples can provide information about the diversity of the SARS-CoV-2 variants and associated mutations at community level.

### SARS-CoV-2 Variants and Mutations of Concern in Europe

During March 2021, most countries in Europe reported an increase in the COVID-19 positive cases. To get an overview of the clinical situation, we determined the count of sequences associated with current VOCs (i.e., B.1.1.7, B.1.351, P.1, and B.1.617.2) (8, 9) in the 20 European countries that had provided wastewater samples (Fig. 1). The increased COVID-19 incidence rate corresponded with the increase in sequences of B.1.1.7 among the clinical samples sequenced in these countries. For P.1 and B.1.351, no general pattern appeared and they were only present to a small fraction. During this time, also very few B.1.617.2 sequences were reported (Fig. 1). While looking at the emergence of the mutations during March 2021, we found that among the most abundant mutations across all countries, the D614G (spike protein) and P323L (non-structural protein, Nsp12) mutations were detected in all the countries (S.fig.2).

**Fig 1:**
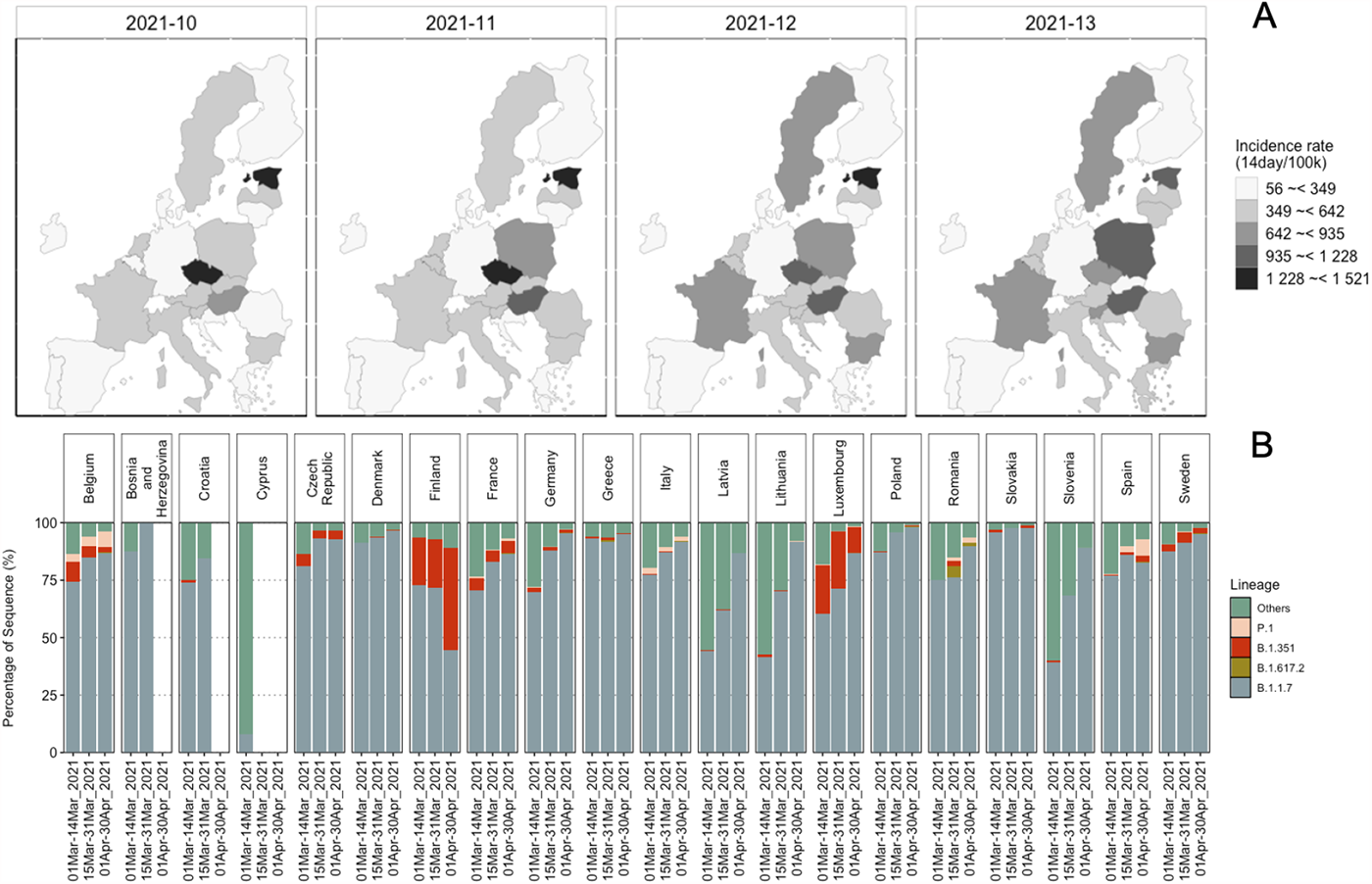
COVID-19 situation in 20 European countries from clinical sequencing data. (A) Maps showing the incidence rate of the COVID-19 positive cases reported in the countries, during weeks 10 to 13, from when the wastewater samples originated. (B) Relative abundance of all sequences for the countries from where wastewater samples were analyzed, available in GISAID (https://www.gisaid.org) on 31-05-2021. All the sequences except B.1.1.7, B.1.351, P.1, and B.1.617 were categorized as “Others”.

Co-occurrences of D614G and P323L have often been reported (24), however, some samples from Germany and China lacked P323L (25). The spike protein N501Y and H69del mutations were also highly abundant in most of the countries, earlier associated with B.1.1.7 and later with all VOCs except B.1.617.2 (S.fig.1, S.fig.2). In some countries the S106del, G107del, F108del mutations, which are ORF1b signature mutations of the VOCs P.1, B.1.1.7 and B.1.351 (26), were also among the abundant mutations (S.fig.2). Across the twenty European countries twenty-six ORF1ab mutations, fourteen spike protein, eight nucleocapsid (N) protein, six ORF8, and three ORF3 mutations were among the dominant mutations, exhibiting spatial and temporal variation (S.fig.2).

### Mutations associated with Variants of Concern in Wastewater Samples

For this study, wastewater influent 24h composite samples from 54 wastewater treatment plants (WWTPs) across 20 European countries were collected between weeks 10 – 13 of 2021 (i.e. 10^th^ to 30^th^ March 2021). We identified the mutations, which vary in their association with the VOCs, in each wastewater sample, by mapping the reads generated to the SARS-CoV-2 reference genome (Wuhan-Hu-1 [GenBank accession numbers NC_045512 and MN908947.3]). In total, 711 different mutations were identified across all the samples, out of which 633 mutations were associated with the VOCs. These 633 VOC associated mutations include mutations that are also associated with other SARS-CoV-2 lineages. Out of these mutations, 619 mutations were observed at >2.5%, 311 mutations at >5% and 23 mutations at >50% allele frequency (Fig.2). Most of the 23 mutations were present in all but a few samples. For example, the W131C mutation was only detected in wastewater samples from Denmark (Fig.2). W131C is one of the important mutations in ORF3a, which is found to assist the ion channel formation and thereby supports the virus in its infectivity (27). The A220V mutation was only identified in samples from Lithuania (Fig.2), which corresponds to the high count of A220V reported in clinical patient samples (S.fig.2).

**Fig 2:**
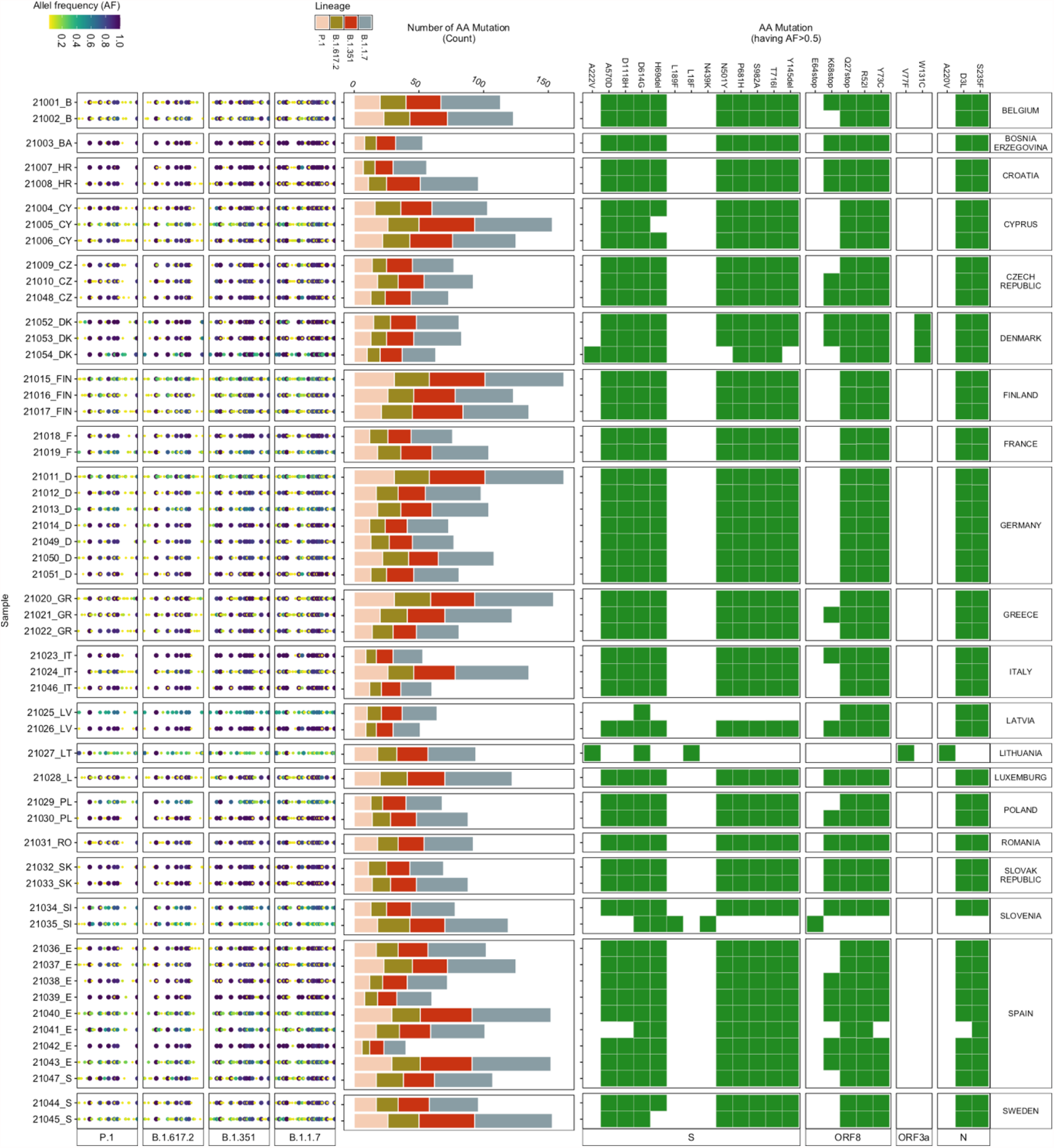
Amino Acid Mutations detected in wastewater samples. (Left) Allele frequency of the mutations (relative to the reference genome Wuhan-Hu-1 [GenBank accession numbers NC_045512 and MN908947.3]), associated with the VOCs, in each wastewater sample. (Middle) Number of mutations (count) detected in each sample corresponding to each VOC. (Right) Heatmap showing the presence (green) / absence (white) of mutations having more than 50 % allele frequency.

Although many low-frequency mutations were observed, the read abundance of low-frequency mutations was mostly similar to the abundance of the high-frequency mutations (S.fig.4). The highest count (ranging between 16-60) was observed for mutations associated with B.1.1.7 in all the samples (Fig.3), followed by B.1.351. P.1 and B.1.617.2, which had a low count of associated mutations but included signature mutations. For example, signature spike protein mutations (i.e. L452R, T478K, P681R, D950N) (9, 28) of B.1.617.2 were identified in some of the wastewater samples.

**Fig 3:**
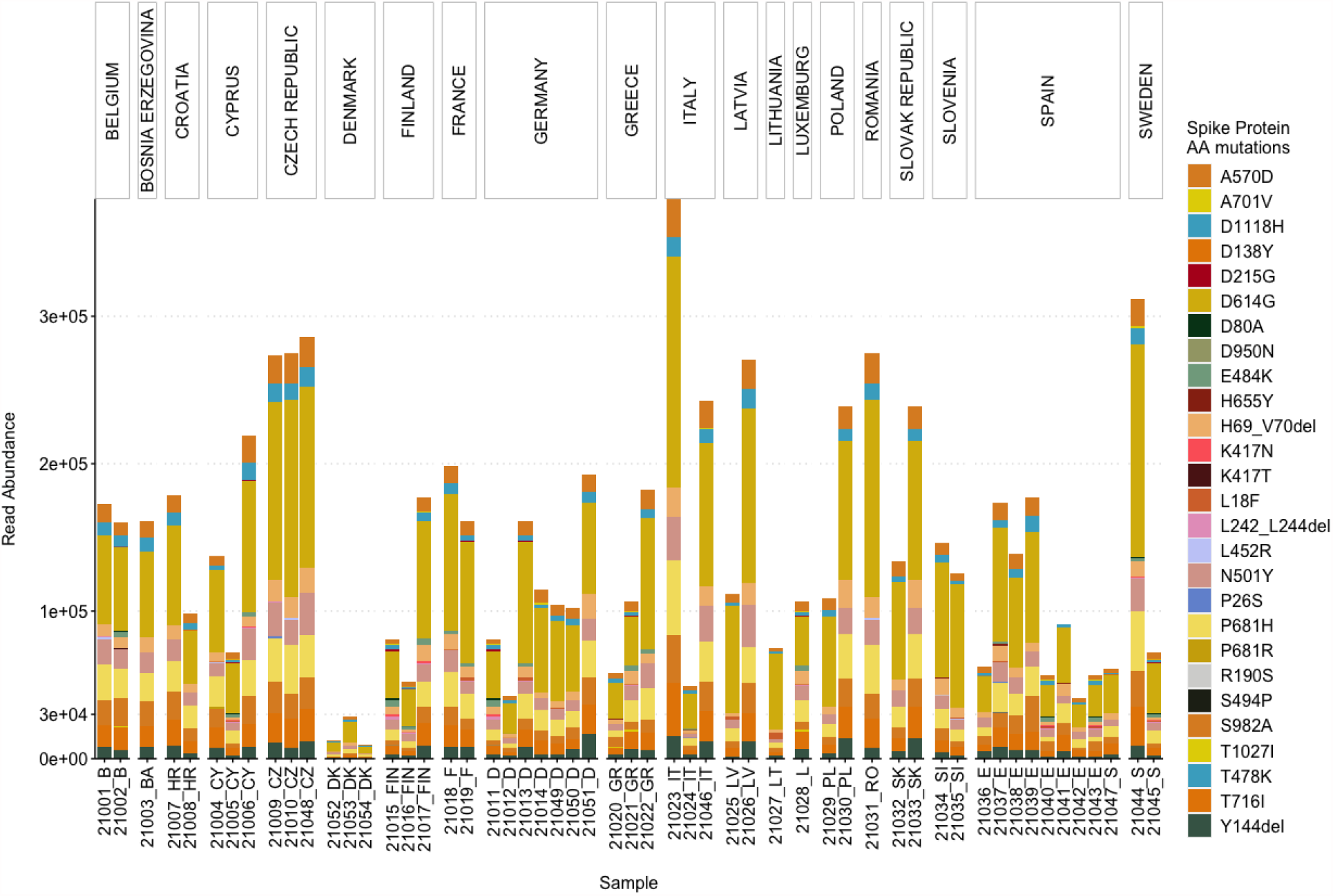
Abundance of the reads associated with Spike protein AA mutations used by the ECDC for the characterization of the VOCs.

### Abundance of Spike protein AA mutations

As the spike protein AA mutations have been associated with changes to characteristics of SARS-CoV-2, leading to an increase in transmissibility and reduced efficacy of treatments, a particular list of spike protein AA mutations is used by the European Centre for Disease Prevention and Control (ECDC) for characterizing the VOCs (9, 29). Therefore, we also assessed the read abundance of these spike protein AA mutations in our wastewater samples (Fig. 3).

Overall, D614G was most abundant, followed by: P681H, T716I, A570D, S982A, H69del, Y144del, D1118H, N501Y, K417N, E484K and others, in decreasing order. Only six out of the 27 AA mutations (i.e. D1118H, D614G, H69del, N501Y, P681H, S982A, and T716I) were present in all the samples (Fig.3). A570D and Y144del were identified in 53 samples. D1118H, S982A, T716I, H69del, P681H and A570D have been mainly found in the B.1.1.7 variant, whereas N501Y has been associated with B.1.1.7, B.1.351, and P.1 (30). For 31 samples, the total read abundance of these AA mutations was above 1e+05 reads, the lowest values in all three sample were detected in the samples from Denmark ranging between 1e+04 to 3e+04 reads (Fig.3).

### Abundance and prevalence of dominant AA mutations

Most of the attention is given to spike protein AA mutations, especially since the emergence of B.1.1.7, because the spike protein mediates virus to host cell-surface attachment, and it is also the principal target of neutralizing antibodies (4). However, mutations in other regions of the SARS-CoV-2 genome are also relevant (30, 31). Therefore, we analysed the most abundant AA mutations found across all the samples for a comprehensive insight into the mutations detected in the wastewater samples (Fig.4). Three mutations (Q27stop, R52I, and Y73C) in ORF8; D3L and S235F in the N protein; P681H, D614G, H69del, S982A, T716I, D1118H, and N501Y in the spike protein; P4804P, T5304T, F3677del, A1708D, S216S, F924F, I2230T, T1001I, F1907F, and H5005H in ORF1ab appeared to be most dominant and prevalent in the samples (Fig.4). The presence of Q27stop and R52I mutations, along with the spike protein mutations, is reported to likely increase the transmissibility of SARS-CoV-2 (32). Another two ORF8 protein mutations, Y73C and K68stop, were present in most of the samples. The Y73C mutation is known to be unique in B.1.1.7 (33). Although no clear relevance of K68stop is known, it is reported to be present in SARS-CoV-2 genomes with the highest number of spike protein mutations (32). The P681H spike protein mutation was the third most abundant across the samples. In total, 13 spike protein mutations were amongst the dominant mutations (Fig. 4).

**Fig 4:**
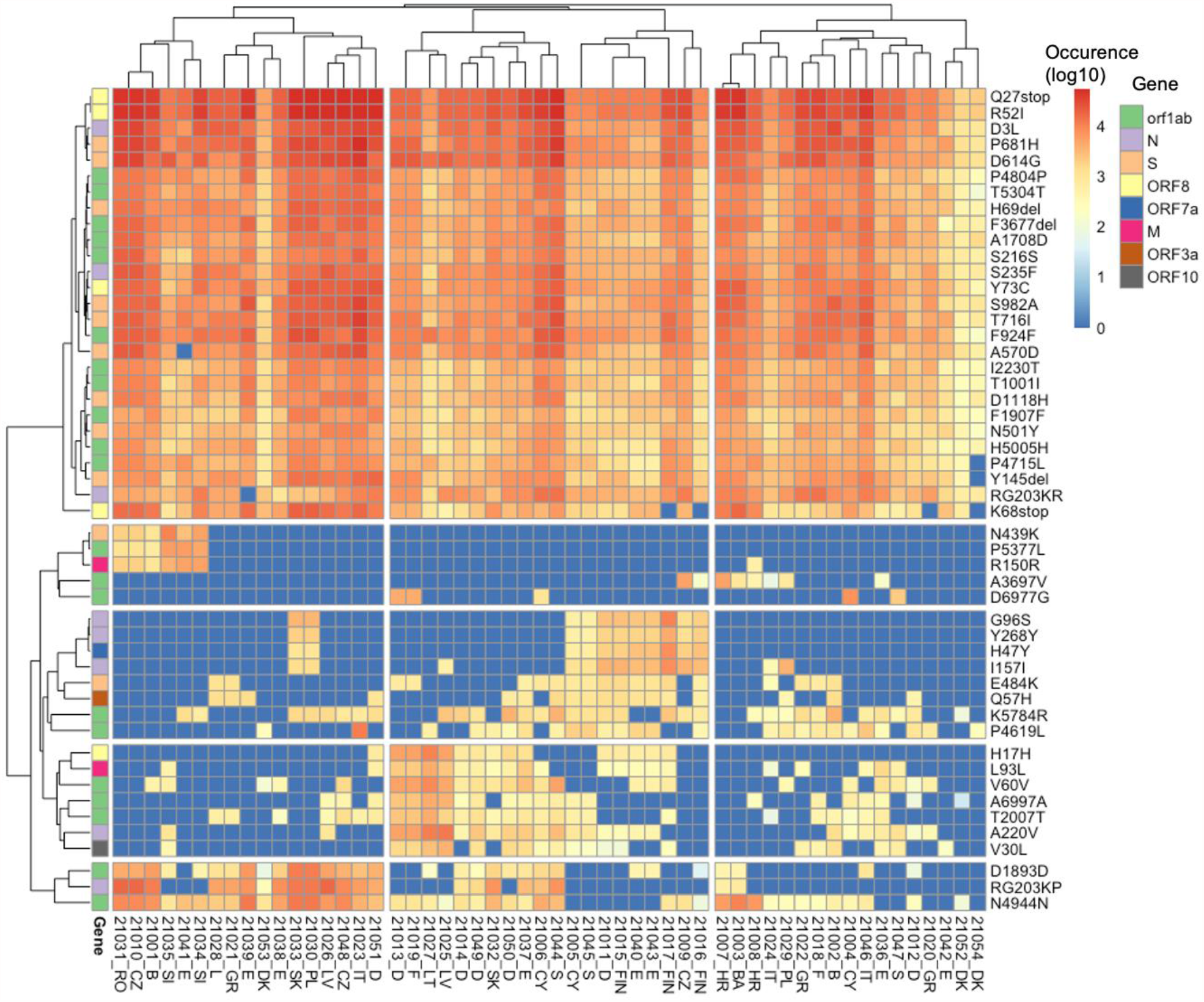
Heatmap showing the read abundance of the top 50 AA mutations found across all the samples.

All spike protein mutations, along with the earlier mentioned ORF8 mutations, have been found in the B.1.1.7 variant in clinical samples (32). E484K was observed in all wastewater samples from France and Sweden. This E484K mutation has been found in B.1.1.7, B.1.351, and P.1(34). Eight mutations of the nucleocapsid protein were also dominant in the samples, especially D3L and S235F. Both of these mutations are known to likely alter the stability and immunogenic properties of the N protein (35) and are signature mutations of B.1.1.7 (36). In the ORF1ab region, 21 mutations were abundant in the samples. The F924F and P4715L ORF1ab mutations have been reported to have a strong allelic association with D614G in variants dominant in Europe (37, 38).

### Wastewater based Epidemiology reveals Genomic Variants

The pattern of genomic variants and the abundance of VOCs were consistent between the clinical and the wastewater sequencing data, especially for dominant mutations (S.fig.2, Fig.4). For example, both data-sets show a high read abundance of the D614G mutation. Similarly, P681H was also dominant in clinical and wastewater samples (S.fig.2, Fig.4). On the other hand, wastewater sequencing data can also reveal genomic variants which are not reported as dominant in clinical data. For example, Q27stop was one of the dominant mutations in wastewater samples but not in the clinical samples of all countries (S.fig.2, Fig.4). The ORF8 mutations (such as Q27stop, R52I, K68stop) have been emphasized to be relevant and require closure attention (32, 39), especially because they occur recurrently (30, 32). However, at present the count of the ORF 8 mutations in genomes deposited in GISAID (https://www.gisaid.org) is low, which according to a previous study is due to bias in samples sequenced (32). Across most of the wastewater samples, we detected a high occurrence of ORF8 mutations (i.e. Q27stop, R52I) (Fig. 4), which provides evidence for the circulation of SARS-COV-2 variants containing these mutations in the sampled regions. Also, wastewater sequencing data can reveal spatial prevalence of mutations. For example, the Q57H mutation was dominant in all three samples from Finland, which is consistent with clinical genomic data of Finland (40). This emphasizes that sequencing for SARS-CoV-2 in wastewater can provide additional information about the prevalence of mutations (such as Q27stop), which might not currently appear abundant based on clinical data. Also relative abundance data grouping the mutations that are associated with a particular variant revealed a clear dominance of B.1.1.7 among the samples, followed by mutations associated with B.1.351, P.1, and B.1.617 (S.fig.5), which is similar to the clinical sequencing data.

The results obtained in this study clearly show that surveillance of SARS-CoV-2 mutation profiles associated with VOCs in wastewater samples is possible using NGS. The data generated also presents the possibility to attain a sufficient coverage of the SARS-CoV-2 genome from the wastewater samples. In wastewater samples, a mixture of genomic material of multiple SARS-CoV-2 variants may be present. Nevertheless, it is still possible to obtain information about variants based on the prevalence of key mutations of the respective variant. However, it is essential to note that sequencing surveillance of wastewater samples should be considered complementary information to whole-genome sequencing of clinical samples.

## Data Availability

All the data is available on NCBI.

## Data Availability

After publication, the data will be made available upon reasonable requests to the corresponding author. A proposal with detailed description of study objectives and the statistical analysis plan will be needed for evaluation of the reasonability of requests. Deidentified data will be provided after approval from the corresponding author and the Mayo Clinic.

## Acknowledgement

We gratefully acknowledge the contribution from the originating laboratories responsible for obtaining the specimens and the submitting laboratories where genetic sequence data were generated and shared via the GISAID Initiative (https://www.gisaid.org). We thank all WWTP operators for providing wastewater samples, We also thank Ray Izquierdo-Lara for critically reading the manuscript.

## Supplementary Material

### Material and Methods

#### Sequencing

For this study, 24 h composite wastewater samples were collected between weeks 10 – 13 of 2021 (i.e. 10^th^ to 30^th^ March 2021) and shipped TU Darmstadt (Darmstadt, Germany), packed with icepacks (approximately at 6°C), for sequencing analysis. In Darmstadt, one litre of the untreated wastewater was filtered through a 0.45 μm electronegative membrane filter to concentrate the SARS-CoV-2 RNA, followed by extraction using the Fast RNA Blue Kit (MP Biomedicals) according to the manufacturer’s protocol. Another 500 ml of the untreated wastewater was concentrated by ultrafiltration in 100 kDa Centricon® Plus-70 centrifugal ultrafilters (Merck) and RNA was extracted using the Ultra Microbiome kit (Thermofisher Scientific) according to the manufacturer’s protocol. Both RNA extracts were pooled together for downstream analysis. From the pooled RNA, cDNA was synthesized using SuperScript(tm) VILO(tm) Master Mix (Thermofisher Scientific), followed by library preparation using the Ion AmpliSeq SARS-CoV-2 Research Panel (Thermofisher Scientific) according to manufacturer’s instructions. This panel consists of 237 primer pairs, resulting in an amplicon length range of 125–275 bp, which cover the near-full genome of SARS-CoV-2. We performed multiple sequencing runs to achieve a high number of reads per sample. For each sequencing run, eight libraries were multiplexed and sequenced using an Ion Torrent 530 chip on an Ion S5 sequencer (Thermofisher Scientific) according to manufacturer’s instructions.

We used the SARS-CoV-2 Research Plug-in Package, which we installed in our Ion Torrent Suite software (v5.12.2) of Ion S5 sequence. We used the SARS_CoV_2_coverageAnalysis (v5.16) plugin, which maps the generated reads to a SARS-CoV-2 reference genome (Wuhan-Hu-1-NC_045512/MN908947.3), using TMAP software included in the Torrent Suite. The summary of mapping of each sample is provided in S.Table 1. For mutation calls, additional Ion Torrent plugins were used, similar to our previous study (1). First, all single nucleotide variants (SNVs) were called using Variant Caller (v5.12.0.4) with “Generic - S5/S5XL (510/520/530) - Somatic - Low Stringency” default parameters. Then, for annotation and determination of the base substitution effect, COVID19AnnotateSnpEff (v1.3.0.2), a plugin developed explicitly for SARS-CoV-2, was used. The raw metagenomic sequence data were uploaded to the National Center for Biotechnology Information (NCBI) Sequence Read Archive (SRA) under Submission ID SUB9829162, BioProject number PRJNA736964.

**S.Table 1:**
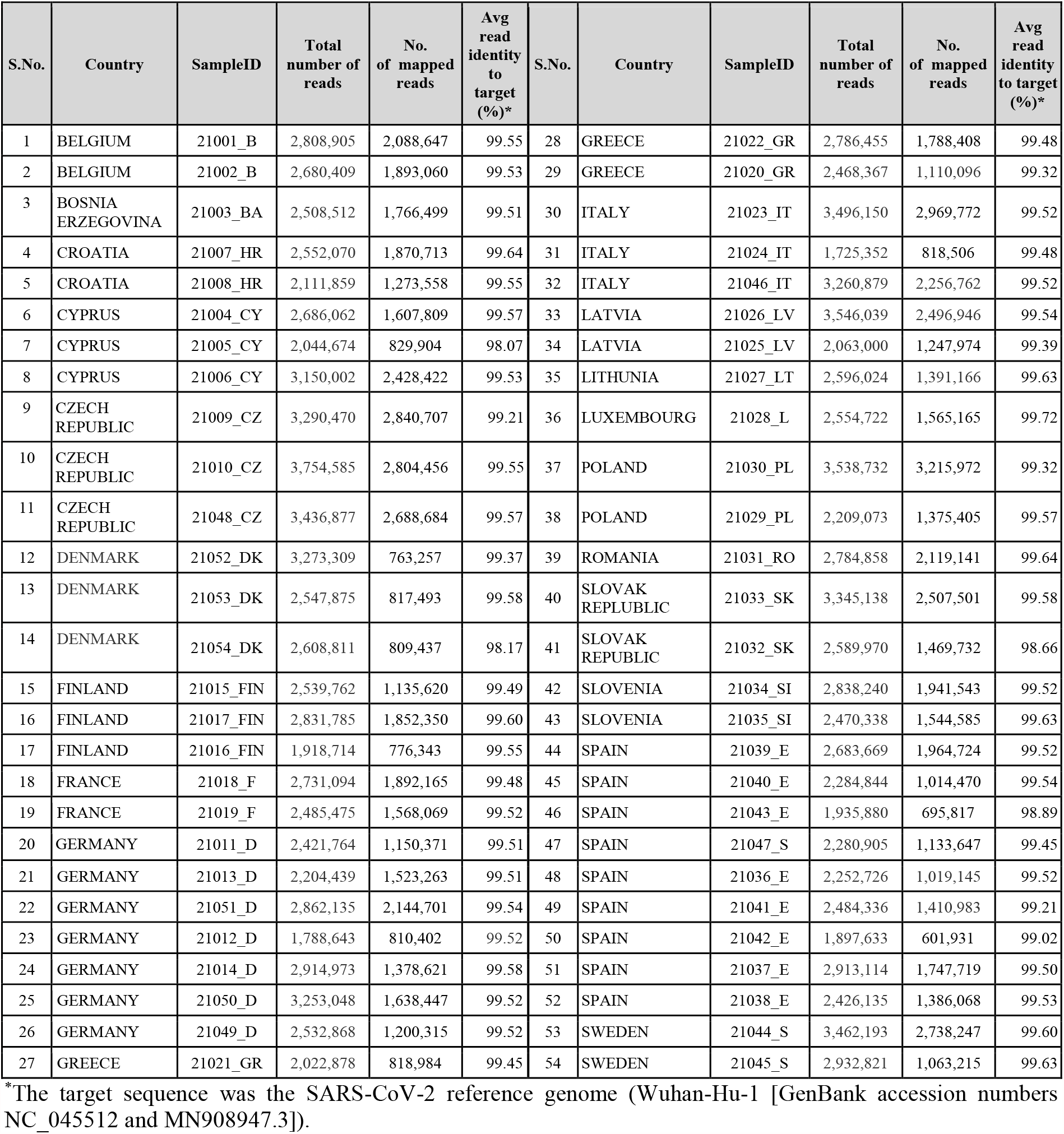
Summary of sequencing data for all samples.

#### qPCR Methods

Samples were received at the KWR laboratory and processed as previously described (2). The N2 assay targeting a fragment of the nucleocapsid gene, as published by US CDC (US-CDC 2020), was used to quantify SARS-CoV-2 RNA in the sewage samples. All RT-PCR’s were run as technical duplicates on 5 µl extracted nucleic acid. RT-qPCR reactions on serial dilutions containing RT-ddPCR calibrated EURM-019 single stranded RNA (provided by the Joint Research Centre) were used to construct calibration curves that subsequently were used to quantify N2 in RNA extracted from the sewage samples. Reactions were considered positive if the cycle threshold was below 40 cycles.

CrAssphage CPQ_064 specific PCR (3) was used to quantify this DNA-virus that is ubiquitously present exclusively occurs in human intestinal tracts in high concentrations. Assays were performed in duplicate on 5 µl 1:10 diluted extracted nucleic acid. Quantification was performed using PCR assays on dilution series of a synthetic quantified gBlock (obtained from IDT, Leuven, Belgium) containing the CPQ_064 gene fragment.

#### Data Analysis

We downloaded the variant surveillance data package from GISAID on 31th May 2021. This data package consists of information about the identified variants, the corresponding amino acid (AA) mutations and sample location. We filtered the dataset, limiting it to human samples with complete coverage. This dataset was used to determine associations between the amino acid (AA) mutations detected in wastewater samples and AA mutations, with their corresponding pangolin lineage, reported from clinical samples. From the GISAID data package, we also determined the fraction of clinical samples reporting the current variants of concern (VOC): (1) B.1.1.7, (2) P.1, (3) B.1.351, (4) and B.1.617.2 (4, 5). Data analysis was performed in R(v3.6.2) using the ggplot (v3.3.3) package for data visualization, and pheatmap (v1.0.12) for hierarchy clustering and heatmap construction.

### Mutations in clinical samples

We determined the abundance spike protein mutations, which are suggested by the European Centre for Disease Prevention and Control (ECDC) for the characterization of the current VOCs (5), reported for clinical human samples. We found that the abundance of the mutations varied for the respective country, though associated with the same VOC (S.fig.1). For example, four out of eight spike protein mutations for P.1 for sequences from Slovakia were not reported.

### SARS-CoV-2 RNA in wastewater samples

#### qPCR results

SARS-CoV-2 N2-gene RNA was detected in all 54 samples in concentrations ranging from 0.4 - 735 gene copies/ml (S. fig 3). The concentration protocol for NGS provided sufficient read depth (S. Table 1), even with the low SARS-CoV-2 concentration samples. The crassphage concentration (S. fig 3) is an index of the dilution of human fecal input in wastewater. This varied between the cities; in INF_21002_B and INF_21003_BA the Crassphage concentrations were markedly low, indicating high dilution of human fecal input in these wastewaters. Normalizing the SARS-CoV-2 N2 concentration for this dilution would markedly change the ranking of the wastewaters, which implies that normalization of ‘raw’ SARS-CoV-2 concentrations in wastewater for the level of dilution of human fecal input is necessary when linking these data to COVID-19 prevalence data.

#### Fraction of variants of concern in wastewater samples

We determined the relative abundance of the VOCs based on the abundance of reads associated with certain AA mutations. As there is quite an overlap between the AA mutations of different SARS-COV-2 variants, for better determination of the relative abundance of the VOCs, we specifically looked for the abundance of unique and shared AA mutations corresponding to each VOC (S.fig.5). Categorization of mutations as unique or shared was based on the percentage of sequences for associated mutations submitted in GISAID. We looked for percentage of sequences for each mutation for every lineage. Then for each VOC, mutations reported in more than 0.5% of total number of sequences for each VOC were selected. Among these selected mutations, mutations that are associated with more than one lineage were categorized as shared mutation otherwise they were associated with the respective VOC. The abundance of mutations associated with B.1.1.7 was highest among the samples ranging from 15 to 40%, followed by abundance of mutations associated with B.1.351, P.1, and B.1.617 (S.fig.5), which is similar to the clinical sequencing data. The mutations associated with B.1.351 were detected in 33 samples, whereas for B.1.617 were detected in 21 samples and for P.1 in 15 samples. B.1.351 mutations were detected in all samples from Finland, Germany, and Sweden; however, the total relative abundance of these mutations varied from 2 to 8%. The relative abundance of the shared AA mutations accounted for 55 to 70% across all samples (S.fig.5).

**S.fig. 1:**
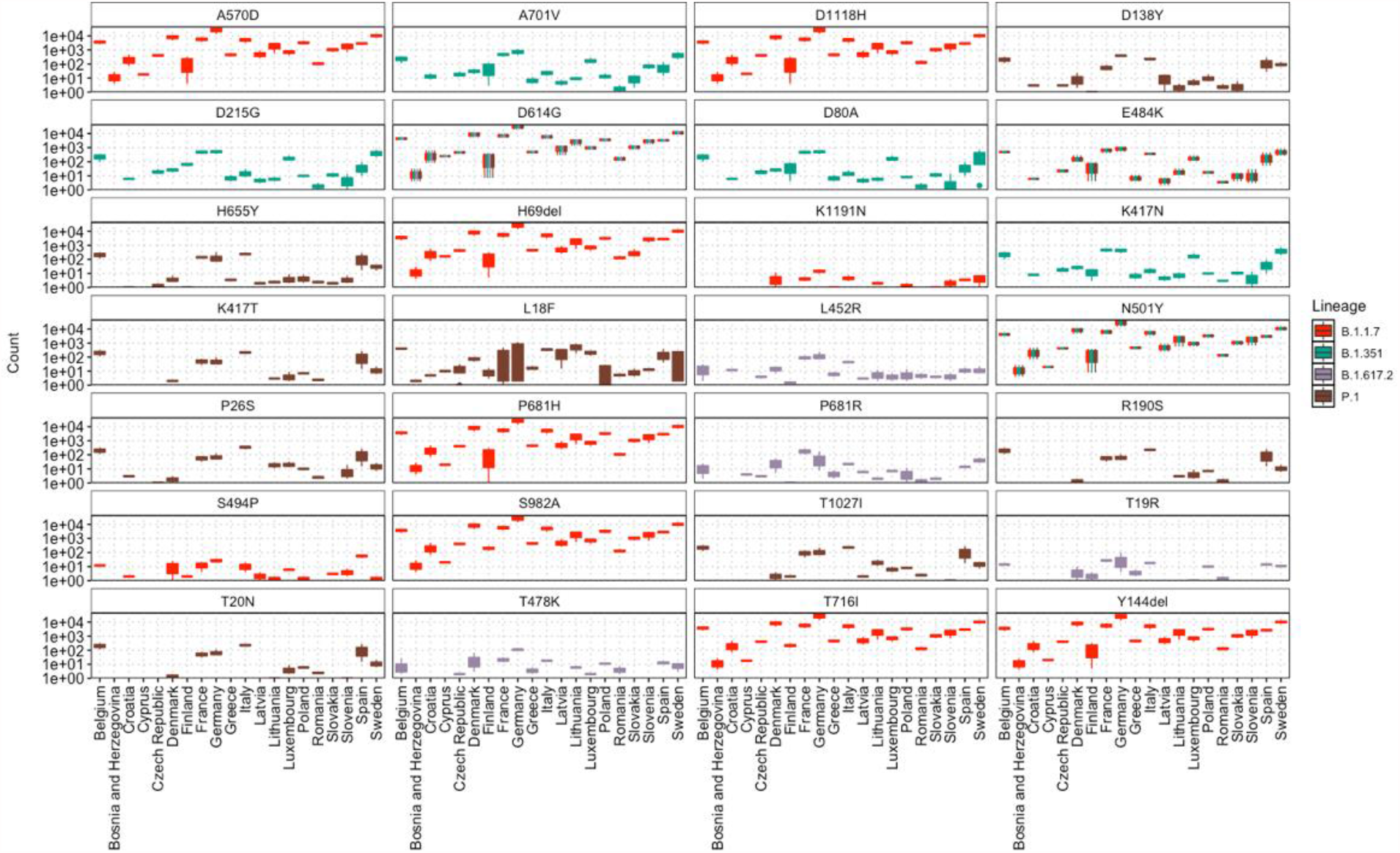
Count (occurrence of mutation in number of genome sequences submitted in GISAID) of spike protein mutations, which are considered for characterization of VOC by ECDC (5,6), found in the variant surveillance data for clinical samples of GISAID dated 31^st^ May 2021. The counts are presented in log10 scale.

**S.fig. 2:**
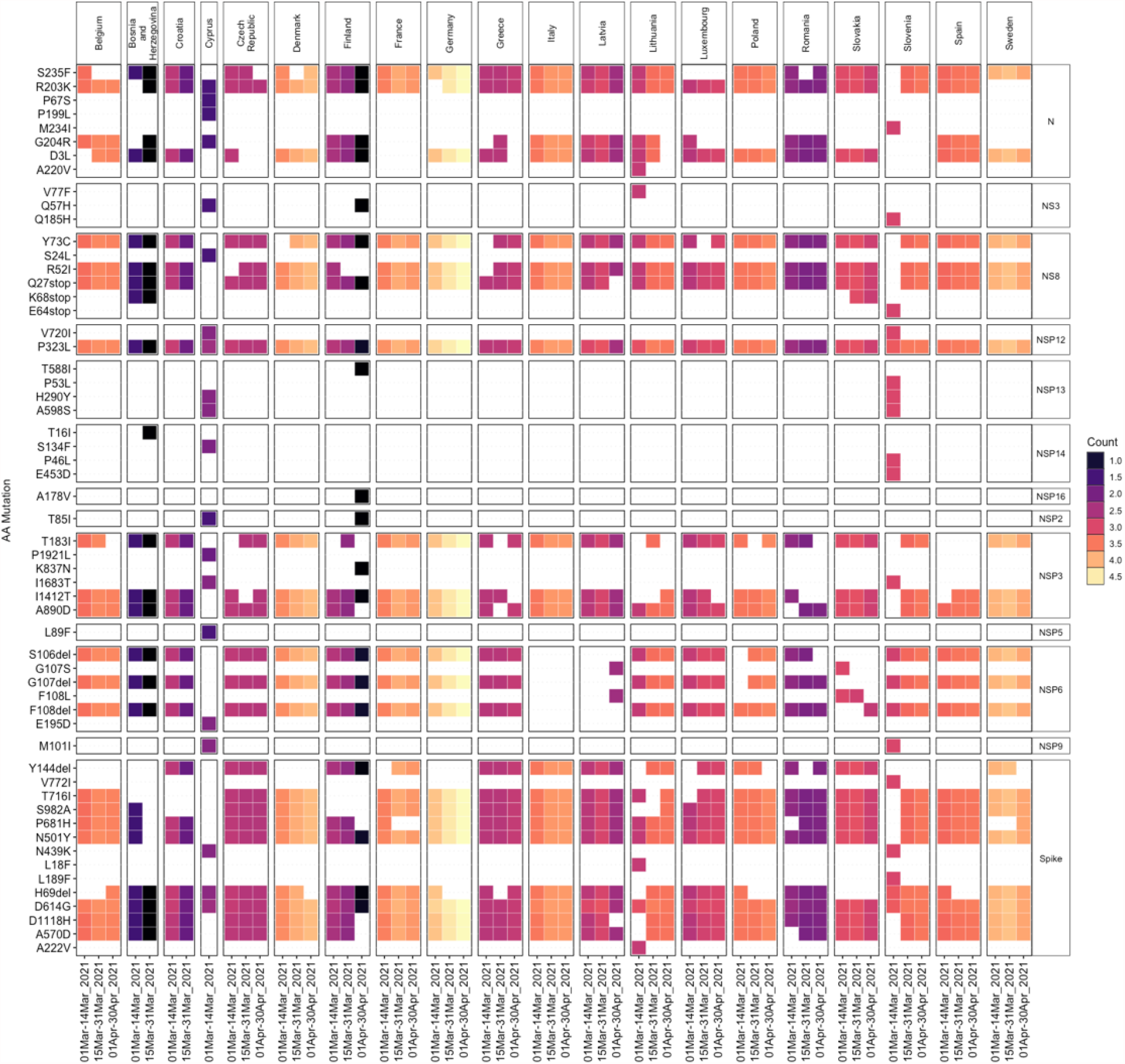
Heatmap representing the top 20 abundant mutations in each country during different time period, based on the count (occurrence of mutation in genome sequences submitted in GISAID) of the mutations.

**S.fig. 3:**
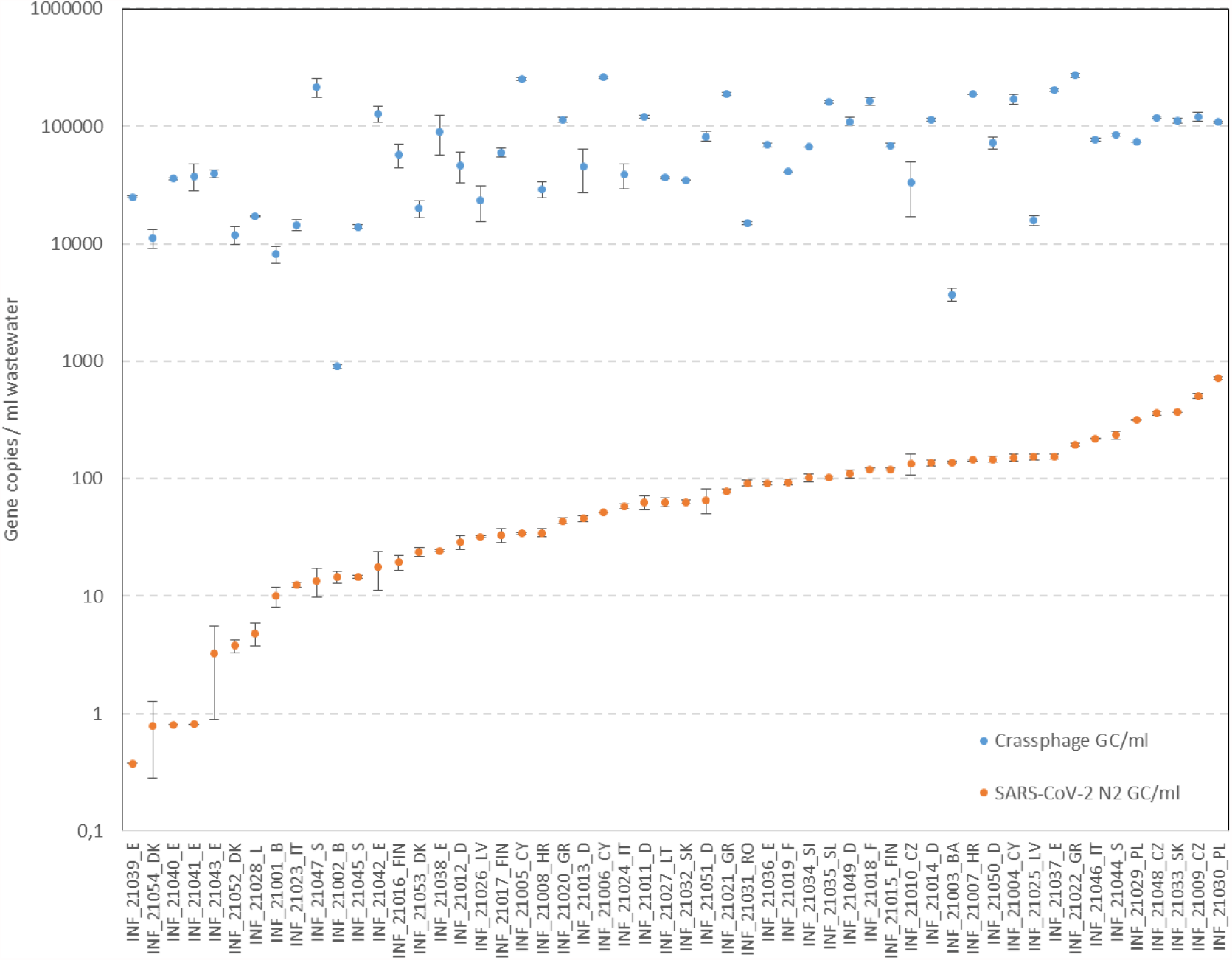
qPCR based analysis showing the concentration of SARS-CoV-2 N2 gene copies detected in each sample.

**S.fig. 4:**
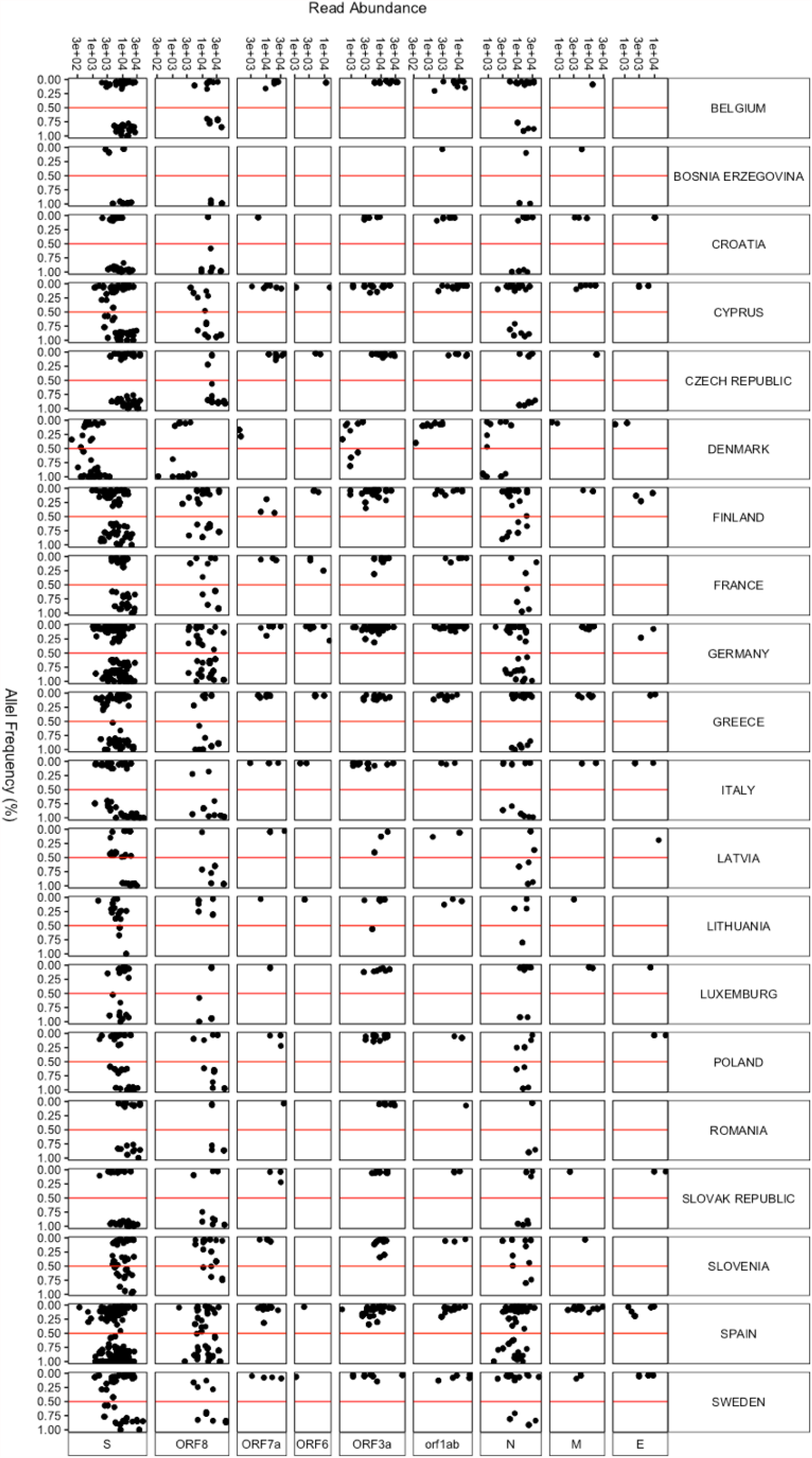
Scatter plot showing the distribution of the read abundance of each mutation against the allele frequency of each mutation.

**S.fig. 5:**
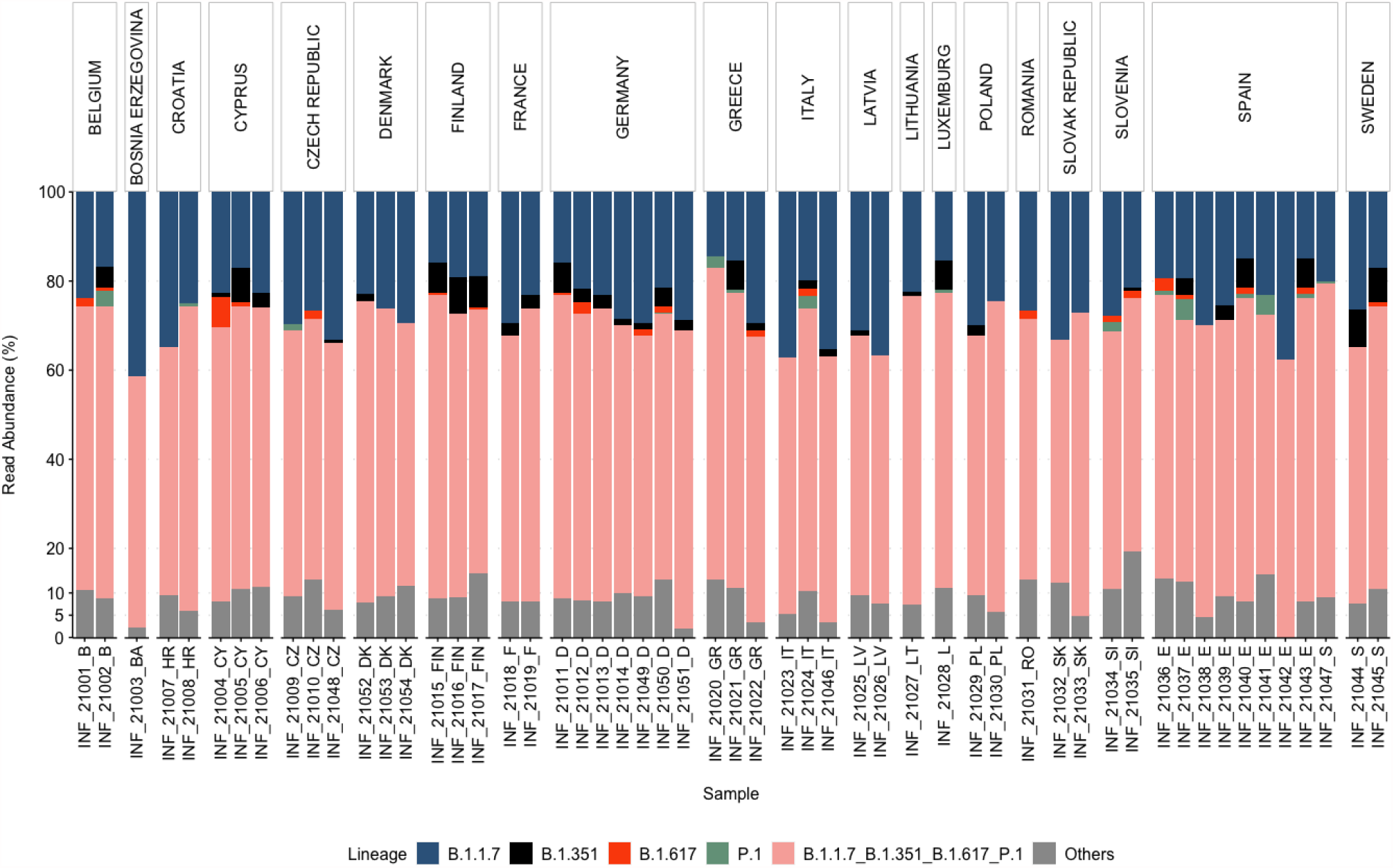
Relative abundance of the variants of concern (VOC) and other variants, based on the abundance of the reads associated with each SNP, respectively. AA mutations shared among different SARS-CoV-2 VOCs are represented as “B.1.1.7_B.1.351_B.1.617_P.1”.

